# Chess-Based Cognitive Remediation Training – A Candidate Add-On-Treatment for Alcohol Use Disorder?

**DOI:** 10.1101/2025.07.29.25332306

**Authors:** Alexandra Seeger, Alexander Kinzel, Kathrin Matthiae, Roland Schmitt, Yury Shevchenko, Sarah Gerhardt, Falk Kiefer, Benjamin Rolland, Juan Antonio Montero, Tillmann Weber, Sabine Vollstädt-Klein

## Abstract

**Aims:** Alcohol use disorder (AUD) is associated with deficits in various cognitive functions which can impair conventional treatment and increase relapse risk. This longitudinal quasi-randomized controlled study investigated chess-based cognitive remediation training (CB-CRT) as add-on therapy to improve cognitive control and psychosocial outcomes compared to standard rehabilitation.

**Materials and methods:** The study was conducted from April 2022 to November 2023 among AUD patients in long-term residential rehabilitation. Participants in the experimental group attended 90-minute CB-CRT group sessions twice weekly for six weeks between April 2022 and April 2023, conducted by clinical staff using the „Entrenamiento cognitivo a través del ajedrez” (ECAM®) method. Assessments were conducted at baseline (T1, day 1), post-intervention (T2, day 42), and on day 126 (T3). Primary outcomes were training effects in sustained attention, decision-making, and set shifting (cognitive flexibility). Abstinence, craving, subjective well-being, and liking of the intervention were assessed at all timepoints (T1, T2, and T3).

**Results:** Fifty-one participants completed two timepoints (T2 six weeks after T1): *n* = 32 allocated to the experimental group and *n* = 19 to the control group (no chess intervention). The CB-CRT group showed significant improvement in sustained attention (*M*_Diff_ = 8.26) and a positive, non-significant trend in cognitive flexibility at T2. No significant effects were found for short-term abstinence, craving, or mood at T3 (day 126). A significant increase in general life satisfaction was observed in the CB-CRT group at T2 (*M*_Diff_ = 3.78), though not sustained at follow-up. The intervention was well accepted, with increased recommendation scores between T1 and later assessments.

**Conclusions:** CB-CRT improved sustained attention and general life satisfaction, and was well accepted. However, the relatively small sample size limited statistical power and detection of further effects. A larger replication study could enhance generalizability and clarify impacts on cognitive control, relapse, and relapse timing.

## 1. Introduction

Alcohol is the most commonly used psychoactive substance in Germany and worldwide and contributes to considerable morbidity and mortality. As stated by the WHO in 2022 [1], alcohol is responsible for an estimated 5.3% of all global deaths and 5.1% of global disability-adjusted life years. Approximately two-thirds of people with an alcohol use disorder (AUD) experience relapse within a year of treatment [2]. Bechara’s neuroanatomical dual process addiction model [3] suggests that an overactive impulsive system impairs the reflective system’s executive control over substance use. This disproportion between impulsive (automatic) and reflective (controlling) systems manifests itself as faulty inhibitory control, which leads to higher risk-taking [3]. As a result, instant rewards, such as the beneficial effects of alcohol use, often take precedence over potential long-term gains, such as the health advantages of sobriety.

Many cognitive and neurobiological addiction theories place executive functions at the centre of the reflective control system [3, 4]. Contemporary addiction theories emphasise the disruption of *executive control* in compulsive drug use [5–8]. *Executive* or *cognitive control*, often used interchangeably with executive functions, encompasses abstract and higher-level constructs such as working memory, attention, problem-solving, and decision-making [9].

Around 50% to 70% of individuals with AUD exhibit neurocognitive deficits compared to healthy controls [10, 11]. These impairments, particularly in executive functions like response inhibition, attention, and working memory, are linked to poorer treatment outcomes [12, 13]. The frontal lobe, crucial for executive control, is especially vulnerable in recently abstinent individuals with AUD, with only limited differences based on the number of prior detoxifications [14, 15]. Despite early cognitive recovery within months of abstinence, a generalized deficit typically persists for several months, with cognitive functions usually stabilizing after one year of abstinence, albeit with some residual impairments [16]. A meta-analysis by Stavro et al. [17] demonstrated that for those significant cognitive deficits, smaller effect sizes are observed following one year of abstinence, indicating progressive recovery over time. However, deficits in visuospatial function may persist even after multiple years of abstinence [17, 18].

*Cognitive Remediation Training* (CRT) is a complementary therapy designed to enhance cognitive functions such as attention, memory, and executive processes, addressing deficits that psychotherapy and pharmacological treatments alone may not resolve [19]. By strengthening cognitive skills, CRT improves treatment outcomes like abstinence and enhances therapies such as Cognitive Behavioral Therapy (CBT) [20]. Metacognitive training may further support individuals with substance use disorders [21, 22]. Enhancing cognition is crucial for improving overall treatment outcomes and social functioning in addiction recovery [23]. More specifically, the early training of executive functions after alcohol detoxification might be crucial in the treatment of AUD, as deficits in higher level executive functions are closely associated with early relapse in this disorder [24].

Though research on CRT’s impact on substance use outcomes is limited, it focuses on experience-dependent recovery through targeted learning strategies and repeated tasks to promote retention and skill transfer [18]. CRT employs both restorative (practicing impaired skills) and compensatory (using alternative strategies) approaches [25]. Tailored programs improve psychosocial functioning, particularly when combined with rehabilitation or social skills training [26]. CRT has shown benefits in conditions like traumatic brain injury, anorexia, and schizophrenia, and recent studies suggest it may enhance executive control and treatment engagement in substance use disorders [27, 28]. While CRT shows promise for AUD in areas such as working memory and inhibitory control [16], findings remain mixed, underscoring the need for further research [28].

Chess has long been studied as a model of expertise and individual cognitive differences [29]. It engages various cognitive functions, such as attention, memory, and problem-solving [30]. Studies highlight its benefits in school education, including enhanced cognitive and academic skills, improved spatial-visual processing, and better pattern recognition [31, 32]. For example, a six-month chess intervention improved meta-cognitive abilities and mathematical problem-solving in schoolboys [33]. Chess players demonstrate superior planning abilities linked to prefrontal cortex activation [34, 35]. In patients with schizophrenia, chess training improved task performance and executive functions, such as task switching and inhibition [36]. Additionally, it can address deficits in substance use disorders, enhancing problem-solving, flexibility, and working memory [37]. Functional magnet resonance imaging studies show that chess stimulates the dorsolateral prefrontal cortex, which is essential for addiction recovery, and improves overall brain connectivity [37, 38]. ‘Motivational chess’, a combination of motivational interviewing and chess training, has shown promise in improving attention, executive functions, and working memory in individuals with cocaine use disorder [39].

Chess-based CRT (CB-CRT) combines traditional CRT with chess training to target cognitive functions such as inhibition, pattern recognition, attention, memory, and planning [40]. Conducted in groups, it uses a chess demo board and incorporates metacognitive strategies like thinking aloud and discussing cognitive processes related to addiction [26, 27]. Participants receive social reinforcement, such as applause, when presenting solutions at the demonstration board, regardless of correctness. This study aimed to evaluate CB-CRT during long-term residential rehabilitation treatment regarding its potential to enhance treatment outcomes for AUD. Based on the theoretical elaboration and previous results, we hypothesised the following: CB-CRT as an add-on intervention improves the test performance in sustained attention, *reward-sensitive* decision-making, *punishment-sensitive* decision-making, and set-shifting in individuals with AUD compared to standard treatment alone. Regarding treatment outcome, we expected CB-CRT to be more efficacious compared to standard treatment regarding abstinence, alcohol craving, mood, and life satisfaction. Finally, CB-CRT was expected to be highly likeable.

## 2. Methods

### 2.1. Study design

Data collection ran from April 2022 until November 2023 at a rehabilitation clinic (MEDIAN clinic Wilhelmsheim, Oppenweiler, Germany). Due to practical constraints, participant assignment to the experimental and control groups was quasi-randomized (see later) rather than strictly randomized as per the study protocol [40]. A single therapist conducted the training on-site during their 15-week stay in the clinic, making smaller group divisions impractical and leading to potential sample size issues. From April 2022 to April 2023, training and experimental group allocation took place at the Wilhelmsheim clinic, while control group assignment occurred mainly toward the end of the chess training period (time-dependent quasi-randomization). Treatment allocation remained concealed until T1. The study design required that participants remained in the clinic for at least six weeks following the scheduling of the T1 assessment, as the chess-based training took place during this period. The T2 assessment was conducted approximately six weeks later, at a time when participants were still in inpatient treatment (see Figure SF1 in the Supplement).

At the time of the follow-up telephone appointments, which took place every 4 weeks (the third follow-up being T3), virtually all participants were no longer in the clinic.

### 2.2. Recruitment

Patients interested in the study were handed out forms regarding their consent to be contacted for the pre-screening phone call, in which the inclusion criteria were queried. The inclusion and exclusion criteria of the study sample, which include MRI criteria as well, are presented in ST1 in the Supplement. A 100€ reward was offered to participants if they completed T1 and T2, as well as an additional amount of 50€ after completing T3.

### 2.3. Intervention

Between T1 and T2, participants in the experimental group attended a 90-minute CB-CRT session twice a week for six weeks. The training was conducted by a clinical staff member using a demo board and exercises of increasing difficulty, focusing on specific cognitive skills such as attention, memory, perception, logical thinking, planning, self-control/impulse control, and decision-making, based on the ECAM® method (https://www.ajedrezmagic.es/metodo-ecam). Exemplary exercises from three trained cognitive domains are displayed in Figure 1.

**Figure 1.**
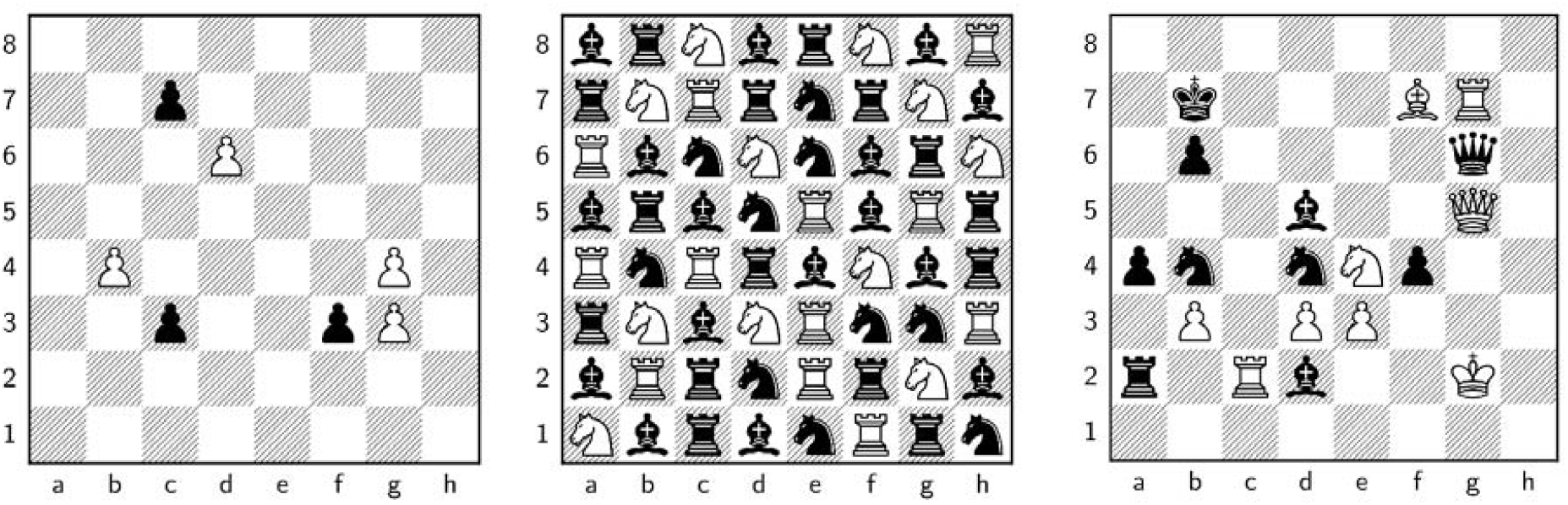
*Note*. **1. Short-time memory**: Participants look at the position for one minute. Then the pieces are removed. Now the participants have to name the squares where the pieces were placed. **2. Sustained Attention**: Participants must mark the white knights on the white squares. The marked knights have to be counted. **3. Executive Functions**: Participants check the legality of the given moves. The vote is yes for legal and no for illegal. Examples: 1. Bishop f7 takes d5 check 2. Knight e4 to c5 check 3. Qg5 takes d5 check.

**Figure 2.**
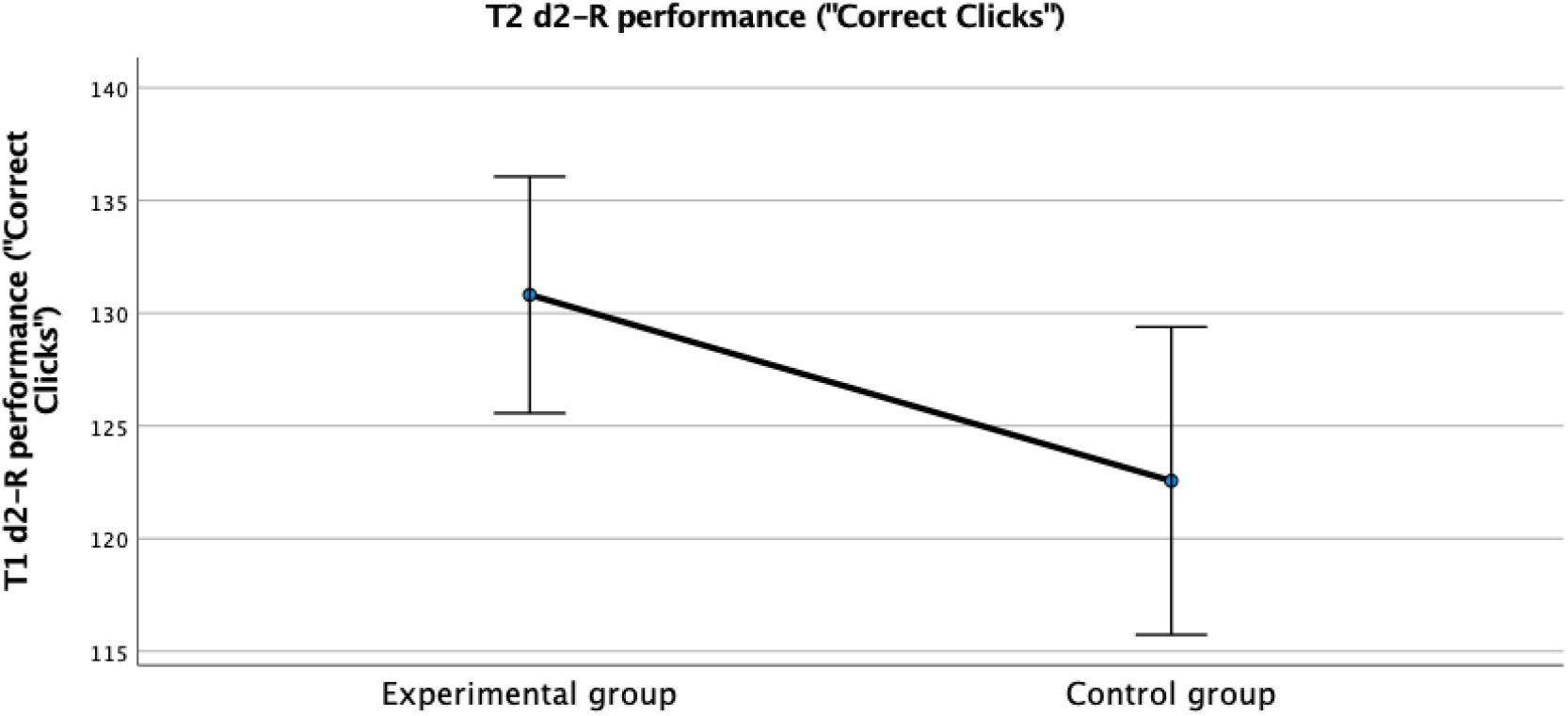
Profile Plot of the adjusted means of the T2 d2-R test performance in both conditions. *Note*. The adjusted mean values of T2 performance (‘correct clicks’), controlling for T1 performance (the covariate), are shown. Error bars are indicated at the 95% *CI*. The overall means for T1 performance (*M* = 114.57; *M(EG)* = 117.81, *M(CG)* = 109.11) is not depicted here.

Completion was operationalised as attending ≥50 % of the 12 CB-CRT sessions (*n*(experimental group) = 32), which corresponds to at least 540 minutes of training. This criterion aligns with the standard length of CBT sessions while accommodating the typical variability in compliance observed in substance abuse treatment settings [41]. As knowledge of the game of chess is not requested but necessary for some of the training exercises, the participants in the experimental group received a handout prior to the intervention in which the set-up of the board and the chess pieces were briefly explained. At the beginning of each CRT session, the instructor explained which cognitive functions were being trained, i.e. psychoeducational information on various concepts of cognitive functions and in particular how these come into play when dealing with craving and help to maintain abstinence. Social reinforcement was also part of the intervention, as everyone clapped when a participant completed a task on the demo board in front of the rest of the group.

### 2.4. Neuropsychological assessment

Each neuropsychological assessment session (test and re-test at T1 and T2) lasted approximately one hour and 45 minutes per participant, iincluding questionnaires on study laptops. Refreshments were provided. The neuropsychological tests included computer-based tasks (∼32 minutes) in the order in which they are outlined, and before, the Letter-Number Sequencing test (∼5 minutes), part of the neuropsychological assessment, was conducted verbally using pen and paper. All computer experiments mentioned below were done via a neurocognitive test battery on the Open Lab online platform [42], which was hosted on an internal server.

**Working memory** was assessed using the Letter-Number Sequencing test (LNS) in its German version by Petermann, which is a subtest of the WAIS-III and WAIS-IV test batteries, used in the age group 16;0 to 69;11 years [43]. The task requires besides attention and concentration the ability for sequential processing, mental rotation, memory span, and auditory short-term memory and assesses cognitive flexibility and fluid intelligence as well [44, 45].

A computerized version of the d2-R test by Brickenkamp et al. [46], a revised and newly standardised version of the d2 attention and concentration cancellation test [47], was used to measure **sustained attention**. The test is used in the age group of 9 to 60 years [46] and measures the ability to concentrate under time pressure, thus also the speed and accuracy in distinguishing similar visual stimuli (detail discrimination) [47]. The outcome variables were correct clicks, error percentages, and accuracy of the test processing, i.e. the number of errors in relation to the number of target objects processed across all trials.

**Decision-making** was operationalized using the Iowa Gambling Task (IGT), which was developed by Bechara et al. [48, 49]. To explore the influence of reward and punishment framing, not only the ABCD version, but also a variant task (EFGH) was included [49]. The instructions emphasized that the ABCD task involved “sometimes losing something” simultaneously, while the EFGH task involved “sometimes gaining something” simultaneously (the order of which task variant came first was randomised between the test subjects). Outcome variables were the total net values summed over all 5 blocks.

The Dimensional Change Card Sort (DCCS) Test was used as a measure of **cognitive flexibility**, often known as task switching, set-shifting, or updating. This task, created by Zelazo and colleagues [50][51], is a shortened version of the Wisconsin Card Sorting Test [52], which assesses cognitive flexibility as part of executive function [53]. Outcome variables were accuracy, reaction time (RT), and a score combining both accuracy and RT using a two-vector scoring method [51]. The cognitive assessments used in this study (d2-R, IGT, DCCS) are established instruments that are suitable for repeated measures, although practice effects cannot be entirely ruled out.

### 2.5. Questionnaires

Sociodemographic data was assessed at T1 by the test performer via REDCap [54], electronic data capture tools hosted at the Central Institute of Mental Health, and included questions on participants’ gender, age, marital status, children, whether they live alone or with another person, highest educational and professional qualification, German language skills, and the diagnosis of mental or neurological illnesses apart from addiction.

All additional questionnaires, except for the Form 90-Interview, were completed by the participants on their own electronic devices using Evasys [55] after each of the assessments had taken place. The Form-90 Interview was conducted before the neuropsychological test battery with the help of a calendar and the data being entered in REDCap through the test performer.

**Recent alcohol consumption** was assessed using Form 90, a standardized calendar-based interview in its German version by Scheurich et al. [56]. For this study, only select variables from the German version of Form 90 were analysed. It was administered at T1 with slightly adapted measures: abstinence time (in days until T1), and alcohol consumption during the 90-day period preceding the last drinking day (quantified in grams and number of drinking days). Additionally, lifetime alcohol consumption (in years) and the corresponding quantity (estimated in standard drinks per day) were assessed at T1. Relapses were assessed at 1-, 2-, and 3-month follow-ups after T2, based on standard drink consumption. These follow-up interviews were conducted via telephone, and responses were documented in REDCap [54].

**Severity of alcohol dependence** was acquired with the Alcohol Dependence Scale (ADS) by Skinner & Allen [57]. **Alcohol craving** was operationalized with a German version of the Alcohol Urge Questionnaire (AUQ) by Bohn et al. [58]. **Subjective well-being** was studied using the Habitual subjective well-being scale (HSWBS) by Dalbert [59], which consists of 13 items, of which six items are assigned to the *mood level scale*, the emotional dimension (originally from Underwood & Froming [60]), and seven items to the *general life satisfaction scale*, the cognitive dimension, from Diener [61].

Finally, we assessed how much the patients enjoyed attending the CB-CRT sessions. For this, we used a self-developed item rated on a point-scale from 1 (not at all) to 10 (very much) in the values for recommending the chess-based training to a friend. It was asked across the different measurement times T1, T2, and T3.

### 2.6. Data analysis

For all analyses, only participants who fully completed the neuropsychological test examinations at T1 and T2 were included. The final raw data of all neuropsychological tasks were pre-processed in R [62]. Sociodemographic and questionnaire data were downloaded from the Evasys and REDCap platforms and imported into SPSS 27.0 [63]. After pre-processing, all analyses (sociodemographic, neuropsychological, and questionnaire data) were done with SPSS. The neurocognitive LNS and the ADS measure are used for descriptive analyses only.

Due to dropout after the examination, the number of participants for the questionnaires can vary as the F90 interview was conducted in person or via phone and the AUQ/HSWBS online.

Linear correlations were conducted using Pearson correlations. The test for normal distribution of relevant variables per group at T1 was carried out using the Shapiro-Wilk test (as *n*(CG) = 19 and not > 30), see ST2 in the Supplement.

To test the effects of the CB-CRT add-on therapy on the 4 neuropsychological variables, one-way analyses of covariance (ANCOVA, independent variable: Condition) were conducted one-tailed at α = 0.05. To adjust for baseline (i.e. T1) differences between the two groups (see ST3 in the Supplement), the T1 measure was included as a covariate and the post-test (i. e. T2) measure was the dependent variable. The strategy was based on average middle-sized underlying Pearson correlations between pre- and post-test results of the whole sample with *r* = 0.5. Questionnaire data measured at 3 timepoints were analysed with a mixed-design analysis of variance (mixed ANOVA), the liking of the CB-CRT intervention with a repeated measures analysis of variance (rANOVA). Events of relapse/dropout were analysed with a Cox regression survival analysis. A sensitivity power analysis was done a posteriori with G*Power 3.1.9.7 [64] for the first hypothesis with the neurocognitive variables “sustained attention (d2-R)”, “decision-making, operationalised separately by the two IGT-versions (IGT-A and IGT-E)” and “mental flexibility (DCCS)”. The power analysis revealed that for a total *N* = 51 of the two groups, at α =.05 and a desired power of (1-β err. prob.) =.80 (an almost 80% probability of finding a training effect, if it exists), an effect size of *f* =.40 can be detected, i.e. moderate to large effects [65].

All group comparisons were tested one-tailed in the hypothesized direction (i.e. improvement by CB-CRT), whereas the Cox regression and ANOVA are inherently not tested one-sided. In the ANCOVAs and mixed ANOVAs, t-tests on the group factor were conducted to test the improvement through CB-CRT one-tailed. All effects are reported as significant at *p* < 0.05.

## 3. Results

### 3.1. Sample Description and baseline data

From initially *N* = 95 screened subjects, *n* = 77 (*EG* = 43, *EG* at T1 = 40; *CG* = 34, *CG* at T1 = 33) were included. Thirty-eight people completed the intervention, but only *n* = 32 participated in the second measurement as did *n* = 19 from the control group (see the Consort Flow Diagram, SF2 in the Supplement). All in all, *n* = 51 subjects that completed T1 as well as T2 were considered eligible for analysis. Of those *n* = 37 (*n* EG = 25) completed all examinations.

Of the 51 participants, 32 people were assigned to the experimental (CB-CRT) group and 19 people to the control group, their ages ranging from 24 to 62 (*M* = 45.67). There was no significant age difference between groups (other sociodemographic characteristics are presented in ST4 in the Supplement). While some descriptive differences (e.g., in level of education) were observed, none reached statistical significance. There were also no significant differences between groups in baseline scores of the AUD-related and psychosocial questionnaires (see ST5 in the Supplement for specific Mann-Whitney U and t-test results). The severity of dependence was moderate (M = 17.77) and did not differ significantly between groups.

### 3.2. Effect of CB-CRT on executive control functions

#### Sustained attention

The mean T2 values adjusted for the T1 values showed a significant higher d2-R test performance in the group with cognitive training (*M* = 130.82, *SE* = 2.61) than in the group without training (*M* = 122.57, *SE* = 3.40), see Figure 4, *F*(1, 48) = 3.67, *p* =.061, two-tailed test, partial η^2^ =.071, when conducting the independent samples t-test on the group factor, *M*_Diff_ = 8.26, 95%-*CI*[-0.41, 16.92], *t*(48) = 1.92, *p* =.0305, Hedge’s *g* = 0.27.

#### Reward- and punishment-sensitive decision-making

There were no significant effects of the treatment group on IGT net values. Adjusted mean net values of the IGT-ABCD at the second measurement time point (post-intervention) in the control group were *M* = 24.55 (*SE* = 4.95) and *M* = 10.86 (*SE* = 3.81) in the experimental group. Consequently, the ANCOVA analysis was not further pursued and the result is considered non-significant. For the IGT-EFGH, the adjusted mean values at the second measurement time point (post-intervention) were *M* = 15.09 (*SE* = 4.43) in the control and *M* = 7.98 (*SE* = 3.41) in the experimental group. Therefore, the ANCOVA was also not pursued for this outcome and is considered non-significant.

#### Cognitive flexibility

Also, no significant effect of treatment was found for cognitive flexibility. The mean DCCS test values adjusted for the T1 were *M* = 9.02 (*SE* = 0.176) in the control group and *M* = 9.06 (*SE* = 0.136) in the experimental group (*M*_Diff_ = 0.04, 95%-*CI*[−0.41, 0.49], *F*(1, 48) =.031, *p* =.86, two-tailed test, partial η^2^ =.001).

### 3.3. CB-CRT Interventional Effect on Treatment Outcome

A Cox regression survival analysis examined the time to relapse. Patients who were not reachable over 3 months from T2 onwards were also classified as relapsers, which is common practice but a more conservative measure. For dropouts, one additional day was added to the survival time. Among the 51 participants, 20 (39.2%) experienced relapse or dropout, while 31 cases (60.8%) remained abstinent/adherent and were censored. The *EG* had a mean of 10.25 standard drinks during relapse (*SD* = 15.21, range: 1–40), compared to 3.5 standard drinks in the control group (*CG*, range: 3–4). Relapse duration also differed, with the *EG* averaging 8.17 days (*SD* = 15.61, range: 1–40), while the *CG* consistently relapsed for only 1 day. The abstinence rates were 62.5% (20/32) in the EG and 57.9% (11/19) in the CG. Overall, the analysis did not find Condition or Abstinence until T1 to be significant predictors of relapse or dropout.

In the Cox regression, the occurrence of the first relapse (together with dropout) was considered as an event, while subsequent relapses and changes in drinking behaviour were addressed using a time-dependent Cox regression. The hazard ratio for the group (*Exp*(B) = 1.134, *p* =.853) suggests a slightly higher risk in the experimental group, but this difference is not statistically significant. However, the hazard ratio for the interaction between group and time (*Exp*(B) = 1.222, *p* =.015) was significant, suggesting that the effect of the group on relapse risk varies over time. Despite this, further analysis, such as recurrent event analysis, could provide even more comprehensive modeling of the relapse process.

The time variable was found to significantly reduce the risk of relapse/dropout (*Exp*(B) = 0.547, *p* =.009), with each unit increase in time associated with a 45.3% reduction in the risk. On the other hand, the duration of abstinence before T1 did not significantly predict relapse/dropout (*Exp*(B) = 0.998, *p* =.646), suggesting that the length of abstinence prior to T1 did not significantly impact on the likelihood of relapse/dropout. The analysis was conducted using two-sided tests and no significant predictors of the risk of relapse or dropout were identified.

### 3.4. Analyses concerning further treatment outcomes

The urge to drink was assessed with the AUQ on 3 examination days in *N* = 33 patients (*n*(*CG*)=8, *n*(*EG*)=25). The two facets of the HSWBS, mood level and life satisfaction, were also assessed on 3 examination days and were analysed in *N* = 35 (*n*(*CG*)=10, *n*(*EG*)=25).

#### Craving

No significant interaction of group x time was found, Huynh-Feldt corrected *F*(1.70, 52.58) = 0.06, *p* =.913, partial η^2^ = 0.002. The main effect of time was not significant, *F*(2, 66) = 0.36, *p* =.668, partial η^2^ = 0.011 and the between-subjects effect of condition was not significant either, *F*(1, 31) = 1.69, *p* = 0.203, partial η^2^ = 0.052, indicating that there was no significant difference of craving between the 3 measurements or the groups.

Adding abstinence time as a covariate also made no difference in the results. For the descriptive statistics at all time points, see ST6 in the Supplement. Both groups had a lower non-significant need to drink at T2 and the EG even showed a greater reduction, without reaching significance. However, if the increase from T1 to T2 was examined by an independent samples t-test, craving for the *EG* is on average 2.38 points lower, 95%-*CI* [-6.04, 1.29]), *t*(45) = -1.31, *p* = 0.099 (tested one-sided), Hedge’s *g* = -0.99.

#### Subjective well-being: Mood and Life satisfaction

In each analysis, there were *n* = 10 from the CG and *n* = 25 from the *EG*. For Factor 1 (Mood level), the interaction time x group was not significant. The **main effect of time** was significant, *F*(2, 66) = 8.85, *p* < 0.001, partial η^2^ = 0.211 (tested two-sided), meaning that there was a small to medium effect difference in Mood Level between the three measurements. The main effect of condition was not significant, see Figure 3. Adding abstinence time as a covariate also made no difference in the results. The descriptive statistics can be found in Table 5 in the Supplement.

**Figure 3.**
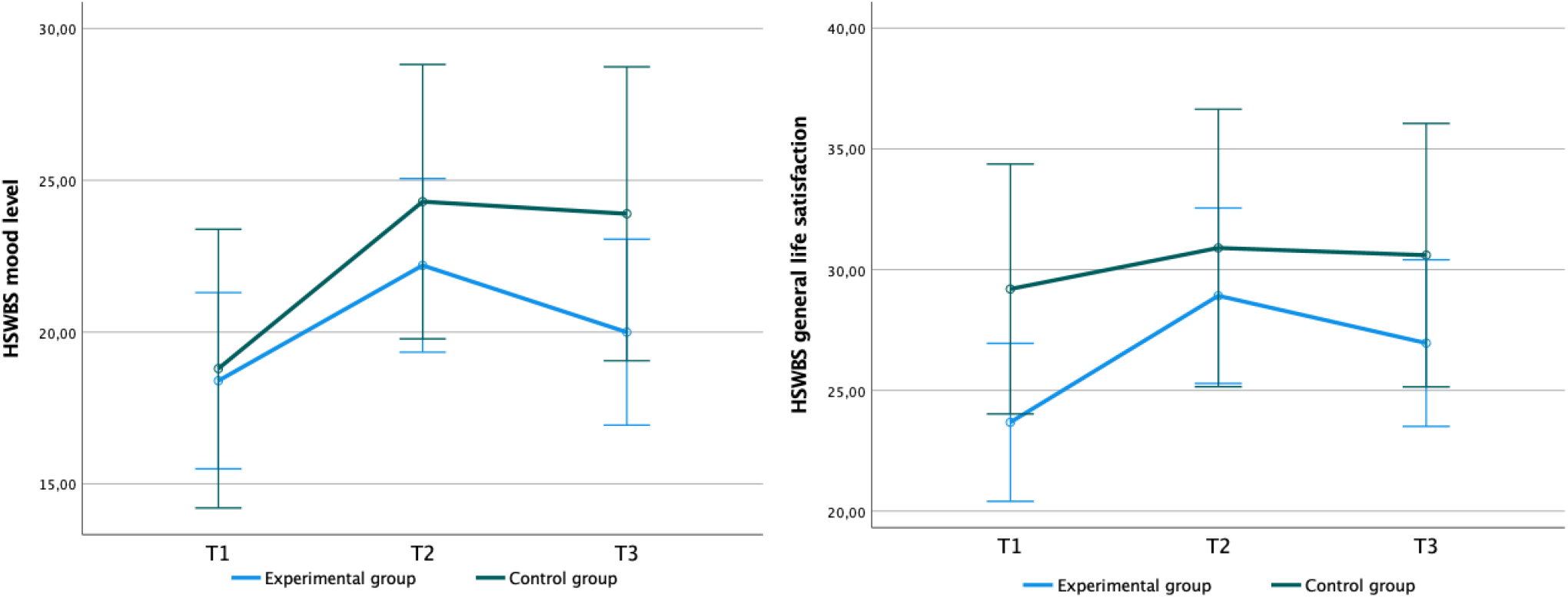
Results of the mixed ANOVAs of Mood Level (left) and General Life Satisfaction (right) *Note*. T1 = Baseline, T2 = After intervention; T3 = 3-month-follow-up after T2. Error bars are indicated at the 95% *CI*.

**Figure 4.**
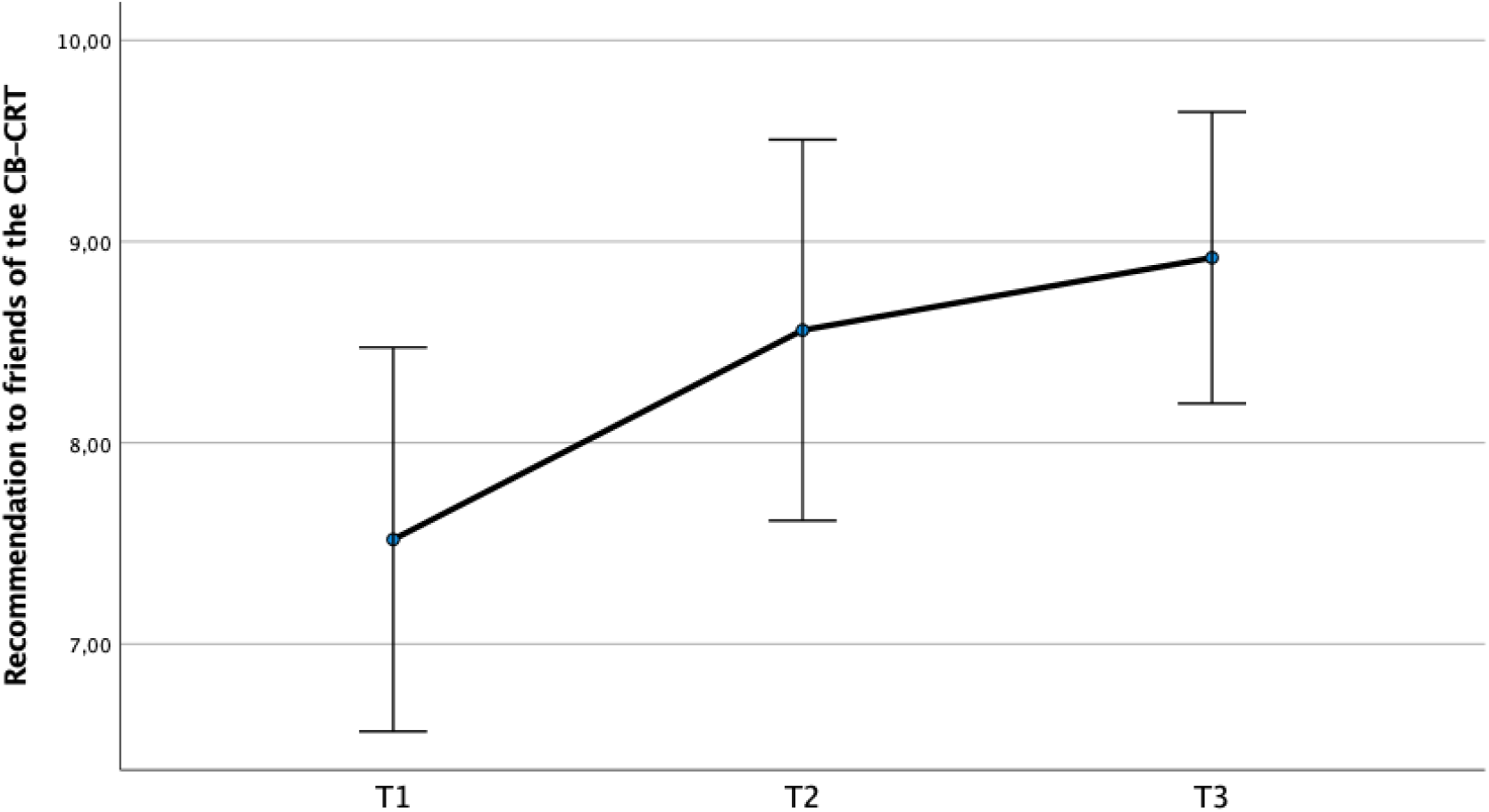
Results of a repeated measures ANOVA of CB-CRT liking in the experimental group. *Note*. T1 = Baseline, T2 = After intervention; T3 = 3-month-follow-up after T2. Error bars are indicated at the 95% *CI*. Experimental group*(n) = 25*. The control group (*n* = 9) is not included here. The question was: “How likely would you be to recommend chess training to a friend?”

For short-term effects in mood level (Factor 1), when considering only T1 and T2, the interaction time x group was not significant. The **main effect of time** was significant, *F*(1, 45) = 14.61, *p* = <.001, partial η^2^ = 0.245 (tested two-sided), indicating a change across both groups, whereas the main effect of condition was not significant.

For Factor 2 (general life satisfaction), the interaction effect between time (T1, T2, T3) and group was not significant. Nevertheless, the within-subjects **effect of time** was significant, *F*(2, 66) = 3.69, *p* =.03, partial η^2^ = 0.101 (tested two-sided), displaying a significant difference in general life satisfaction between the three measurements. However, there was no effect of condition (group), see Figure 3. Adding abstinence time as a covariate also made no difference in the results. For descriptive statistics, see Table 5 in the Supplement.

To examine short-term effects in general life satisfaction (factor 2), only T1 and T2 were included in the model. In this analysis, the **interaction effect** time x group was significant, *F*(1, 45) = 3.26, *p* =.078, partial η^2^ = 0.068 (tested two-sided). The **main effect of time** was also significant, *F*(1, 45) = 7.01, *p* =.011, partial η^2^ = 0.135 (tested two-sided), whereas the main effect of condition was not significant. The interaction effect was driven by a larger life satisfaction increase in the EG from T1 to T2 (independent samples t-test: life satisfaction for the *EG* is on average 3.78 points higher (95%-*CI* [-0.44, 8.01]), *t*(45) = 1.81, *p* = 0.039 (tested one-sided), Hedge’s *g* = 0.54).

#### Liking of the CB-CRT intervention

The experimental group (*n* = 25) was asked if they would suggest the CB-CRT intervention to a friend (recommendation from 1 (not at all) to 10 (very much)). There was a significant **main effect of time** for the recommendation values across the different measurement times (*F*(2, 48) = 8.25, *p* <.005, partial η^2^ =.256). These changes were significant between T1 (*M* = 7.52, *SD* = 2.31) and T2 (*M* = 8.56, *SD* = 2.29), *p* <.05, and between T1 and T3 (*M* = 8.92, *SD* = 1.75), *p* <.001, according to the calculation of contrasts, all tested two-tailed. There was no significant increase in mean scores between T2 and T3 (*p* = 0.339), see Figure 6. The results suggest that likeability scores changed over time with a large effect according to Cohen [67], with significant differences found between T1 and the later time points, while the increase between T2 and T3 was not significant.

## 4. Discussion

Given that improvements in cognition are commonly observed during abstinence and the associated ‘time-dependent recovery’ of cognitive functions [68], the present study aimed to examine whether participation in chess-based cognitive remediation training (CB-CRT) could promote additional improvements beyond standard rehabilitation treatment. Specifically, we investigated whether CB-CRT would enhance executive control functions (sustained attention, decision-making, and cognitive flexibility) and improve psychosocial outcomes (abstinence, craving, subjective well-being, and liking of CB-CRT) relative to a control group receiving standard rehabilitation without CB-CRT. Therefore, findings are presented as improvements in the experimental group relative to the control group.

### Training Improvements of Executive Control Functions

The d2-R-task is a commonly used experiment for testing sustained attention, selective attention, and concentration [46]. The results indicate that chess-based CRT improves performance in the d2-R-task. The main effect of condition (partial η^2^ =.071) is a medium effect according to the classification of Cohen [65]. Although the two-tailed confidence interval [-0.41, 16.92] includes zero, the one-tailed test and Hedge’s *g* = 0.27 support a significant effect with small to medium effect size [65] in the expected direction. Thus, CB-CRT appears effective for erhancing sustained attention, including selective attention and concentration.

No significant group differences were found for decision-making abilities in either the reward- or punishment-sensitive versions of the Iowa Gambling Task. A possible explanation is that some participants may have misunderstood the instructions. Alternatively, decision-making, as a complex problem-solving function [69], may be more resistant to rehabilitation, particularly when involving hot executive functions. Another factor could be that the control group had, on average, a slightly longer (though non-significant) abstinence period, which may have benefitted their cognitive recovery at T2, in line with time-dependent cognitive recovery theories [18]. However, for cognitive flexibility, the CB-CRT group showed a slightly higher but non-significant improvement after treatment, as measured by the DCCS test. Given that the ANCOVAs achieved a power of.80 and could only detect medium-sized effects (f = 0.40), smaller effects might have remained undetected. It is also possible that shifting is less impaired in chronic alcohol use disorder compared to substance use disorders more broadly [12]. Moreover, shifting’s relevance for self-regulation may be context-dependent [70], with acute alcohol consumption only moderately impairing shifting [71], but latent shifting abilities showing alcohol-induced deficits, especially in individuals with initially lower baseline shifting skills [72]. Notably, to date, few studies have investigated cognitive training effects on shifting, with the exception of our pilot CB-CRT study [40], which found improvements in trail-making flexibility after 14 weeks in a substance use disorder population.

### Intervention Effect on Treatment Outcome

An unfavorable treatment outcome was defined as “relapse or drop-out” in the Cox regression, as drop-out can hypothetically indicate an undetected relapse. Results suggest that participants in the experimental group were more likely to relapse or drop out than those in the control group; however, the difference on this combined measure was not statistically significant. Notably, the higher dropout rate alone in the control group points to poorer treatment retention and lower adherence to the study protocol. Although not statistically significant, these findings may indicate a trend worth exploring in future studies regarding abstinence and treatment retention, as well as the potential benefits of CBT-based strategies for maintaining long-term abstinence. Given the higher dropout rate in the control group, opting for an active control condition instead of a ‘treatment-as-usual’ group might be more advantageous.

### Additional analyses about further treatment outcomes

The results suggest that the CB-CRT did not result in distinct changes in craving and mood level. Further, there were some observable trends, such as an initial reduction in craving and an initial increase in (habitual) mood. The lack of significant effects implies that the chess intervention did not have a substantial impact on these psychosocial factors, also when controlled for abstinence time. Future research could explore other confounding factors that may influence these outcomes, as this may seem to depend on intraindividual factors, such as the patient’s social network. On the other hand, there were higher craving ratings in the experimental group (*M* Diff = 2.61) from the beginning (T1), despite only very small differences in mean Alcohol Dependence Scale (ADS) scores (*M* Diff = 0.31, *EG* > *CG*). Higher craving could be related to more pronounced bodily perceptions and interoceptions, as in the Interoceptive Awareness System [73] rather than reflecting the actual severity of AUD. As the CB-CRT has been shown to improve cognitive control functions, it could have potentially helped balance out these interoceptions over time with an even longer commitment. This third component to the dual-process models leads people to occasionally sense temptations, i.e., conscious momentarily salient desires to engage in a rewarding behaviour as substance use, modulating the balance between impulsive and reflective system, as impairment of this third system can eliminate such craving desires [74]. However general life satisfaction was increased in the CB-CRT group at T2, but not persisting until the 3-month follow-up. While Cibeira et al. (2021) observed significant improvements in the quality of life within the chess-training group in their pilot study [75], this study found significant between-group differences, indicating a stronger impact on life satisfaction. These differences may reflect variations in sample characteristics (older adults in Cibeira’s study), but both studies suggest that similar interventions can improve quality of life [75], with the effectiveness potentially influenced by participant age and the focus on subjective well-being.

### Liking of the CB-CRT intervention

The high initial likeability score (*M* = 7.52/10) indicated that participants found the CB-CRT recommendable even before starting. Likeability further increased significantly from pre-to post-intervention and from pre-intervention to the 3-month follow-up, accompanied by a large effect size. The average adherence rate within the experimental group was 84.6% (*SD* = 11.81; *Med* = 83.3%), ranging from 50% to 100%. Most participants attended over 80% of sessions, with 15.6% completing all 12 sessions, though some variability in participation was observed.

Overall, the high likeability and strong adherence highlight participant commitment. The increase in satisfaction over time may reflect the motivational structure and accessibility of the training. Given that CB-CRT requires only a demonstration board, pieces, and simple staff instructions, its clinical applicability appears promising. These findings underline the potential for broader implementation of the CB-CRT.

### Limitations

This study represents a first step in evaluating chess-based CRT for AUD but is not without limitations. Due to its exploratory nature and limited sample size, multiple comparison corrections were not applied, increasing the risk of Type I errors; future research should address this with appropriate statistical adjustments. The relatively small sample size, while acceptable for a clinical longitudinal study, limited statistical power and the ability to detect smaller effects. A replication study with α = 0.05, 1-β =.95, and *f* =.25 would require more than twice the current sample size to improve generalizability and clarify effects on sustained attention, cognitive flexibility, short-term abstinence, dropout rates, and life satisfaction.

Although the Cox regression did not find significant group effects on relapse or dropout risk, recurrent event analysis could offer more comprehensive modeling of the relapse process. A possible explanation for the limited transfer effects is the specificity of the training: apart from improved sustained attention, there was only a minimal, non-significant improvement in cognitive flexibility and no effect on decision-making. This aligns with Caetano et al. [76], who criticized cognitive training in substance use disorders for often being too narrow to generalize. Future programs should incorporate a broader range of cognitive targets. Moreover, more complex cognitive functions, which are harder to rehabilitate [69], should be examined separately from simpler ones.

No significant differences in education level were found between groups, suggesting that selection bias is unlikely. Nevertheless, future studies should assess participants’ chess knowledge more systematically or provide standardized pre-training to ensure equal baseline familiarity. Additionally, the lack of significant effects on mood, craving, and short-term abstinence may also be explained by the fact that both groups received intensive relapse prevention treatment as part of standard care, combined with generally low relapse rates across the sample. Although the control group showed slightly better short-term abstinence outcomes, it also exhibited higher dropout rates, underscoring the complexity of therapy retention dynamics.

## 4.1. Summary

This research confirms some of the findings of our pilot study, as cited in the study protocol [40], and extends them by including a control group, examining relapse likelihood combined with dropout after CB-CRT, and assessing psychosocial effects using objective measures.

Although the generality of the current findings needs to be established by future research, the present study has supported a moderate improvement in sustained attention through CB-CRT. The results show only minor non-significant effects on abstinence and reduced dropout rates, when combined. Additional analyses do not indicate effects on craving and mood. However, subjective general life satisfaction improved post-intervention, although this effect did not persist at the three-month follow-up. Notably, the intervention was well received, as indicated by a significant increase in likeability ratings over 3 time points.

Taken together, despite some limitations, this research represents a first step towards the evaluation of chess-based CRT for AUD in a clinical trial. Although the generalisability of the current findings needs to be established by future research, the present study has provided some support for an improvement in general life satisfaction, in sustained attention and by trend in cognitive flexibility through chess-based CRT as well as a likeability of the intervention, which therefore can be classified as possible candidates for adjunctive treatment in inpatient settings. It can also be described as a strategy-based CRT approach that emphasises planning, pattern recognition and memory through engaging exercises based on the game of chess. Future research should explore specific AUD subgroups, such as those with varying levels of cognitive impairment, or extend findings to inpatient or ambulatory settings with patients having longer abstinence and partially rehabilitated cognitive functions. It remains to be investigated whether a longer-term intervention could improve life satisfaction in the longer term.

Recent guidelines from a Delphi consensus study recommend delivering cognitive remediation training multiple times per week for over three months, focussing on impaired cognitive functions identified through individualised neuropsychological assessments [77]. This approach is more frequent and tailored than the CB-CRT examined in this study. A promising digital development is the training app GYMCHESS© (https://gymchess.com), which implements the CB-CRT exercises, ensuring variety and appropriate difficulty.

## Data Availability

The data that support the findings of this study are available from the corresponding author upon reasonable request.

## Ethics and dissemination

This study was embedded in the clinical fMRI trial for alcohol as well as tobacco use disorder. The trial has been approved by the Ethics Committee of the Medical Faculty Mannheim at the University of Heidelberg (reference number 2017-647N-MA). Before study inclusion and after a detailed explanation of all procedures, all participants have given written informed consent. The study was registered in the Clinical Trials Register (trial identifier: NCT04057534) on December 8th, 2019.

## Funding

This study was supported by a grant from the Deutsche Forschungsgemeinschaft (Project ID 421888313). The Deutsche Forschungsgemeinschaft was not involved in the planning of the study, in data collection, analyses or publication procedures.

## Acknowledgements

We would like to thank Carmen Gaitan Coronado and Ainoa Jiménez Recuero for their training in ECAM®. Gereon Lex and Jennifer Holzammer helped with study initiation. Simon Kaminski conducted the CB-CRT. Heiner Fritz and Xenia Schalasta helped with measurements. Besides SVK, Edgar Erdfelder was a supervisor and Beatrice G. Kuhlmann co-supervisor of the first author’s master thesis on which this manuscript is based on. Damian Karl and Alfred Wieland helped with the draft of a manual on ECAM®.

## Authors contributions

SVK and TW designed the study. SG and SVK supported the implementation and data collection of study. JAM developed ECAM®, on which CB-CRT is based. RS, JAM and SVK adapted the training. AS wrote the manuscript and analysed the data. RS carried out the CB-CRT as an instructor. AK, AS and KM collected the data. YS programmed the neuropsychological test battery and supported data analysis. All authors (AS, AK, KM, RS, YS, SG, FK, BJ, JAM, TW, SVK) were involved in the interpretation of data and in revising the draft of the manuscript critically for important intellectual content and read and approved the final manuscript.

## Conflicts of interest

All authors have no conflict of interest to declare.

## Supplementary Material

The supplementary material includes detailed inclusion and exclusion criteria (Table 1), additional statistical analyses such as Shapiro-Wilk testing, skewness, and descriptives (Table 2), descriptive data on neuropsychological measures at T1 (Table 3), demographic information (Table 4), and results from group comparisons on AUD-related and psychosocial questionnaires (Table 5). It also contains a summary of AUQ and HSWBS characteristics (Table 6), the study procedure (Figure 1), and a CONSORT flow diagram (Figure 2).

## Notes

### Competing Interest Statement

The authors have declared no competing interest.

### Clinical Trial

NCT04057534

### Clinical Protocols

https://bmjopen.bmj.com/content/bmjopen/12/9/e057707.full.pdf?fbclid=IwAR0x-fQvhG8X1s7nChH6Q0fNYPrm6EKP0eH3Q55C-XMdtcnSE9Wm535DB5s

### Author Declarations

This study was embedded in the clinical fMRI trial for alcohol as well as tobacco use disorder. The trial has been approved by the Ethics Committee of the Medical Faculty Mannheim at the University of Heidelberg (reference number 2017-647N-MA). Before study inclusion and after a detailed explanation of all procedures, all participants have given written informed consent.

