## Supplementary Material for "Chess-Based Cognitive Remediation Training – A Candidate Add-On-Treatment for Alcohol Use Disorder?"

**Tables**

**ST1**

*Inclusion and Exclusion Criteria of the Study Sample*

| Inclusion criteria | Exclusion criteria |
| --- | --- |
| • Age between 18 and 65 years  • Good knowledge of German  • Normal or corrected to normal vision  • Ability to consent to study (no legal guardian): Signed written informed consent and signed consent for data security  • Alcohol use disorder according to DSM-5 (more than three fulfilled criteria)  • Currently in therapy for AUD (in the rehabilitation clinic for > 10 weeks)  • Abstinence from alcohol >72 hours | • Positive alcohol test  • Common exclusion criteria for MRI (e.g., pregnancy, metals in the body, claustrophobia, epilepsy, adiposity)  • Suicidality or endangerment of others  • Severe cognitive impairments (e.g., dementia)  • Severe physical illness  • Severe neurological disorders, history of brain injury  • Therapy with methylphenidate within the last 8 weeks  • Other mental disorders, except for mild or moderate anxiety, phobias, adaptation, post-traumatic stress, personality, attention deficit/hyperactivity disorders |
|  | • Other Axis I mental disorder except for mild, moderate or remitted depression, other substance use disorders if AUD is still the main diagnosis  • Severe withdrawal symptoms or intoxication  • Psychotropic medication within the last 14 days except for antidepressants or soporific and intake of medication for treating withdrawal effects until 3 days prior to study participation |

*Note. DSM = Diagnostic and Statistical Manual of Mental Disorders. MRI = Magnetic resonance imaging.* Adapted from our study protocol [40].

**ST2**

*Results of Shapiro-Wilk Testing and Descriptives of Neuropsychology test variables, AUD-related and psychosocial variables at T1*

| Shapiro-Wilk-  Test |  | CB-CRT group | |  |  | Control group | |  |
| --- | --- | --- | --- | --- | --- | --- | --- | --- |
|  | *n* | *M* | *SD* | *p (skewness)* | *n* | *M* | *SD* | *p (skewness)* |
| DCCS Performance | 32 | 8.88 | 1.03 | **.005** (neg.) | 19 | 8.77 | 1.23 | **.002** (neg.) |
| D2-R Performance | 32 | 117.81 | 27.98 | .834(sym.) | 19 | 109.11 | 25.11 | .675(sym.) |
| IGT-A Net Score | 32 | 2.5 | 17.98 | .735(sym.) | 19 | -1.26 | 25.94 | .1(sym.) |
| IGT-E Net Score | 32 | 8.94 | 23.26 | **.001** (pos.) | 19 | 5.16 | 20.84 | **.006** (pos.) |
| LNS Sum Score | 31 | 21.77 | 3.08 | **.006** (pos.) | 18 | 21.44 | 3.42 | .712(sym.) |
| Years of heavy alcohol | 31 | 8.79 | 8.67 | **.00** (pos.) | 19 | 10.05 | 8.36 | **.02** (pos.) |
| Standard drinks p.d. in those years | 31 | 57.84 | 66.96 | **.00** (pos.) | 19 | 72.42 | 102.37 | **.00** (pos.) |
| Abstinence | 32 | 83.84 | 63.13 | **.00** (pos.) | 19 | 126.16 | 104.66 | **.001** (pos.) |
| ADS Sum Score | 32 | 17.92 | 6.53 | .13 (pos.) | 18 | 17.61 | 7.02 | **.05** (pos.) |
| AUQ Sum Score | 32 | 13.78 | 7.24 | **.00** (pos.) | 19 | 11.68 | 4.40 | **.001** (pos.) |
| HSWBS mood level | 31 | 19.13 | 7.61 | .401 (pos.) | 19 | 20.84 | 7.40 | .892 (pos.) |
| HSWBS general life satisfaction | 31 | 24.68 | 8.43 | .338 (pos.) | 19 | 28.58 | 8.41 | .643 (pos.) |

*Note.* Abstinence in days until T1. All measures are indicated as for T1. Significant p-values are highlighted in bold. Positive Skewness indicates a right-skewed distribution.

**ST3**

*Descriptive statistics of the neuropsychological measures for the whole sample at T1*

| Neurocognitive Measures, *N* = 51 |  | |  | |  |
| --- | --- | --- | --- | --- | --- |
|  | *Min.* | *Max.* | *M* | *St.Err.* | *SD* |
| **D2-R performance (correct clicks)** | 52 | 169 | 114.57 | 3.78 | 27.03 |
| BZO | 81 | 291 | 144.63 | 5.11 | 36.52 |
| AF | 1 | 199 | 30.06 | 5.76 | 41.17 |
| VF | 0 | 5 | 0.75 | 0.18 | 1.26 |
| F% | 0.59 | 81.27 | 18.63 | 2.53 | 18.09 |
| **DCCS performance** | 5.53 | 10 | 8.84 | 0.15 | 1.09 |
| Correct responses | 51 | 60 | 58.53 | 0.30 | 2.17 |
| Median reaction time in ms | 379.8 | 1038.23 | 596.43 | 20.66 | 147.54 |
| Reaction time score | 0.86 | 4.14 | 3.96 | 0.15 | 1.07 |
| Accuracy | 4.25 | 5 | 4.88 | 0.03 | 0.18 |
| **IGT-A total net score** | -60 | 60 | 1.10 | 2.96 | 21.12 |
| Block 5 net score | -20 | 20 | 1.29 | 0.97 | 6.94 |
| **IGT-E total net score** | -24 | 60 | 7.53 | 3.12 | 22.26 |
| Block 5 net score | -20 | 20 | 2.63 | 1.31 | 9.34 |
| **LNS sum score ^a^** | 16 | 30 | 21.65 | 0.45 | 3.18 |
| LNS process value **^a^** | 4 | 8 | 6.14 | 0.17 | 1.21 |

*Note.* ^a^ *N* = 48. D2-R = Revised d2 attention and concentration cancellation test. DCCS = Dimensional Change Card Sort (test). IGT = Iowa Gambling Task. LNS = Letter-Number Sequencing test.

**ST4**

*Sociodemographic Characteristics of Participants at T1*

| Baseline characteristic | CB-CRT group | | Control group | | Full sample | |
| --- | --- | --- | --- | --- | --- | --- |
|  | *n* | % | *n* | % | *n* | % |
| Gender |  |  |  |  |  |  |
| Female | 3 | 9.4 | 4 | 21.1 | 7 | 13.7 |
| Male | 29 | 90.6 | 15 | 78.9 | 44 | 86.3 |
| Marital status |  |  |  |  |  |  |
| Single | 13 | 40.6 | 6 | 31.6 | 19 | 37.3 |
| Married/partnered | 13 | 40.6 | 6 | 31.6 | 19 | 37.3 |
| Divorced/widowed | 6 | 18.8 | 7 | 36.8 | 13 | 25.5 |
| Children ^a^ | 18 | 56.3 | 14 | 73.7 | 32 | 62.7 |
| Cohabitating | 17 | 53.1 | 10 | 52.6 | 27 | 52.9 |
| Highest educational level |  |  |  |  |  |  |
| Volks-/Hauptschule | 7 | 21.9 | 9 | 47.4 | 16 | 31.4 |
| Realschule | 13 | 40.6 | 7 | 36.8 | 20 | 39.2 |
| (Fach-)Abitur | 12 | 37.5 | 3 | 15.8 | 15 | 29.4 |
| Highest job level |  |  |  |  |  |  |
| No qualification | 2 | 6.3 | 2 | 10.5 | 4 | 7.8 |
| Ausbildung/Lehre | 24 | 75 | 17 | 89.5 | 41 | 80.4 |
| Fachhochschule | 1 | 3.1 | 0 | 0 | 1 | 2.0 |
| Hochschule | 5 | 15.6 | 0 | 0 | 5 | 9.8 |
| Mother tongue German ^a^ | 27 | 84.4 | 15 | 78.9 | 42 | 82.4 |
| Mental (as well as minor neurological) illness besides addiction ^a^ | 9  (3) | 28.1  (9.4) | 9  (0) | 47.4 (0) | 18 (3) | 35.3  (5.9) |

*Note. N* = 51 (*n* = 32 (EG), *n* = 19 (CG)). Percentages refer either to the respective condition or the total sample, as indicated.

^a^ Reflects the number and percentage of participants answering “yes” to this question. The highest educational and job level categories remain in the German wording to maintain comprehensibility.

**ST5**

*Results of Mann-Whitney-U and independent-samples t-tests examining AUD-related and psychosocial questionnaire parameters at T1*

| Questionnaire data | | *U* Z | | *t*(48) | *p* | Pearsons’s *r* /  Hedges’ *g* |
| --- | --- | --- | --- | --- | --- | --- |
| **Mann-Whitney U-tests**  1. Years of heavy alcohol | 254.00 | | -0.81 | - | .417 | -0.12 |
| 2. Standard drinks p.d. in those years | | 270.50 | -0.48 | - | .631 | -0.07 |
| 3. Abstinence until T1 | | 219.00 | -1.66 | - | .098 | -0.23 |
| 4. ADS | | 270.00 | -0.36 | - | .716 | -0.05 |
| 5. AUQ | | 247.00 | -1.13 | - | .260 | -0.16 |
| **Independent-samples t-tests**  6. HSWBS Mood Level | | - | - | -0.78 | .439 | -0.22 |
| 7. HSWBS General Life Satisfaction | | - | - | -1.59 | .118 | -0.46 |

*Note.* ADS = Alcohol Dependence Scale, AUQ = Alcohol Urge Questionnaire, HSWBS = Habitual subjective well-being. *n*(EG) = 31 for measures 1, 2, 6, and 7; 32 for the other measures. *n*(CG) = 18 for measure 4; 19 for the other measures. The group differences in *N* were taken into account.

**ST6**

*Descriptive Characteristics of the AUQ and HSWBS (2 factors), as of the mixed ANOVAs*

|  |  | CB-CRT group | |  | Control group | |  | Full sample | |
| --- | --- | --- | --- | --- | --- | --- | --- | --- | --- |
|  | *N* | M | SD | *N* | *M* | SD | *N* | *M* | SD |
| **AUQ (Craving)** |  |  |  |  |  |  |  |  |  |
| T1 | 25 | 13.36 | 7.30 | 8 | 10.75 | 3.99 | 33 | 12.73 | 6.69 |
| T2  T3 | 25  25 | 11.56  12.92 | 4.02  7.96 | 8  8 | 10  10.5 | 3.51  2.98 | 33  33 | 11.18  12.33 | 3.91  7.11 |
| **HSWBS Mood Level** |  |  |  |  |  |  |  |  |  |
| T1 | 25 | 18.4 | 7.29 | 10 | 18.8 | 6.70 | 35 | 18.51 | 7.03 |
| T2 | 25 | 22.2 | 6.98 | 10 | 24.3 | 7.15 | 35 | 22.8 | 6.99 |
| T3 | 25 | 20 | 7.04 | 10 | 23.9 | 8.69 | 35 | 21.11 | 7.63 |
| **HSWBS General Life Satisfaction** |  |  |  |  |  |  |  |  |  |
| T1 | 25 | 23.68 | 8.1 | 10 | 29.2 | 7.87 | 35 | 25.26 | 8.31 |
| T2 | 25 | 28.92 | 8.46 | 10 | 30.9 | 10.08 | 35 | 29.49 | 8.84 |
| T3 | 25 | 26.96 | 8.52 | 10 | 30.6 | 8.53 | 35 | 28 | 8.52 |

*Note. N* = 35 (*n* = 25 (EG), *n* = 10 (CG)).

**Figures**

**SF1**

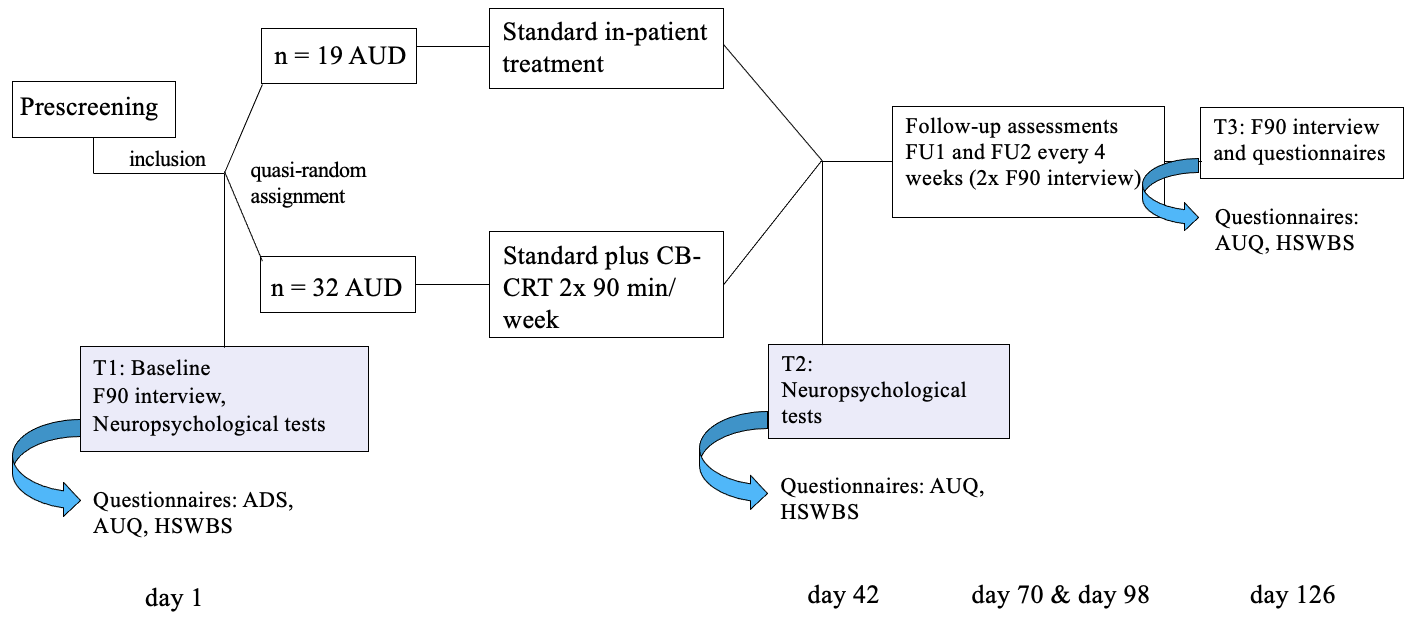
*Study procedure*

*Note.* ADS = Alcohol Dependence Scale, AUQ = Alcohol Urge Questionnaire, HSWBS = Habitual subjective well-being. The questionnaires (including the Form 90 Interview) and the neuropsychological test battery are explained afterwards. Exact number of days between assessments varied slightly.

**SF2**

*Consort flow diagram*
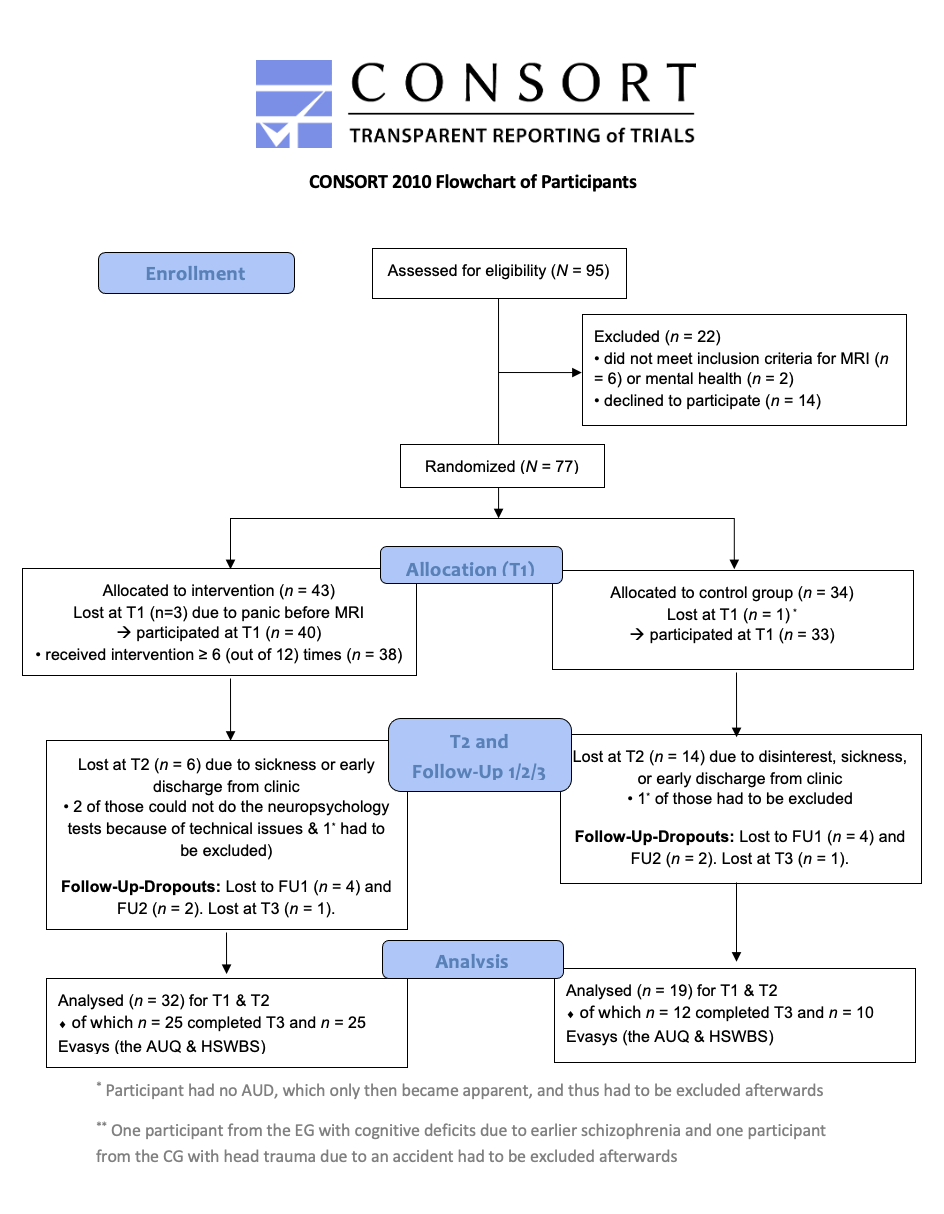
